# Descriptive analysis of the Autism Spectrum Quotient (AQ) in a sample of Brazilian adults

**DOI:** 10.1101/2020.08.04.20168179

**Authors:** Ana Luíza Costa Alves, Jonas Jardim de Paula, Débora Marques de Miranda, Marco Aurélio Romano-Silva

## Abstract

**Background:** The Autism Spectrum Disorder is characterized by the presence of difficulties in social interaction, inflexible, repetitive and/or stereotyped behaviors and interests. The Autism Spectrum Quotient (AQ) is a self-report instrument with 50 items created to quantify autistic traits in individuals older than 18 years with average or above-average intelligence.

**Objectives:** The principal aim was to present a brief descriptive analysis of the AQ in a heterogeneous sample of Brazilian adults, also, to measure the clinical validity of the scale.

**Method:** We recruited all the participants in Brazil (N=1024). Then we described the distribution in the general population (N=385) and investigated the AQ accuracy in a sample of autistic adults (N=32).

**Results:** Our results suggested that autism traits were normally distributed in the population, but Brazilian adults have shown a different profile from the original study. Further, we found that 24 adults from the sample had a clinical score on the AQ, compatible with their previous autism diagnosis.

**Discussion:** Our population probably reports more symptoms compared to other because the original clinical score represents a lower percentile in our sample. Also, future work will be required to adequate the use of the AQ in the Brazilian population.

## INTRODUCTION

People with Autism Spectrum Disorder (ASD) show difficulties in two main areas: social interaction, including verbal and non-verbal communication deficits, and inflexible, repetitive and stereotyped behaviors and interests (APA, 2013). These traits could be perceived in the early development, the diagnosis is currently made around the age of 3 years, although some behaviors can emerge as early as during the first year. Those symptoms impact functionality in different domains, including relationships, professional life, academic outputs, and mental health (Purpura et al., 2020). ASD is a heterogeneous condition seen as a spectrum, where people may differ in how intense the symptoms are, which may range from mild to severe presentation. This aspect will determine the impairments and the kind of support the person will need.

The prevalence of ASD around the world and across all ages is approximately 1% (APA, 2013) but evidence also suggests that autism traits show a normal distribution in the typical population (Constantino and Todd, 2003). Major difficulties with social behavior, which is a core symptom of autism, could be common in people with no such diagnosis (Constantino and Todd, 2003; Rutter, 2011). The assessment of ASD in childhood is well documented in the literature with many adapted and validated clinical instruments. However, this scenario is different when we consider adults with ASD, especially when the clinical condition is less severe, with average intelligence and no delay in language development.

With the proposal to quantify ASD traits in individuals older than 18 years old with IQ in the normal range, Baron-Cohen et al. (2001) developed the Autism Spectrum Quotient (AQ), which is a self-report questionnaire with 50 items that are divided into five different domains: social skills, imagination, communication, attention switching, and attention to details. The cut-off score can identify the number of symptoms the individual present, classifying for severity, and for the need and kind of support necessary. Although the diagnosis of autism should be made by a team of multidisciplinary professionals, the use of instruments and questionnaires have the objective to give support to the diagnostic process. Thus, the AQ scale enables health professionals to identify some important traits of autism in the adult population. The AQ was adapted for many cultures, including for the Brazilian Portuguese by Egito et al. (2018).

As the clinical importance of this instrument and the normal distribution of those traits in the general population are known, we aimed to briefly present a descriptive analysis of AQ scores in a sample of Brazilian adults without autism diagnosis or any other psychiatric condition, followed by validation of its accuracy in a sample of adults with ASD.

## METHODS

This study was approved by the Research Ethics Committee of the Federal University of Minas Gerais-UFMG and the consent was obtained from all volunteers before the study. The research was organized in different stages. In the first one, we adopted an online platform for data collection. Subjects were invited by direct mailing and in social media, especially targeting groups of researchers which work with autism and groups of autism patients and relatives, along with other people who would like to volunteer. We received 1024 valid form submissions in our server. A series of exclusion criteria were applied in this initial sample: age below 18 years, self-reported history of mental disorders or neurological diseases, use of psychotropic medication and scores above the cut-off score for mental disorders (>7) in the Self Reporting Questionnaire-20 (SRQ-20) (Mari & Williams, 1985). The final sample of this study involved 385 individual (294 women), with mean age of 34.3 years (SD=11.3, range=18-68).

Then, we invited part of the sample who got a clinical score (>31, the international cutoff) on the AQ or reported a previous diagnosis of ASD to a diagnostic interview, following the American Psychiatric Association ‘s Diagnostic and Statistical Manual of Mental Disorders (DSM-V) criteria. Only participants which lived in Belo Horizonte-MG and were interested in the procedure, as reported in our online form were invited. This subsample was formed by 32 voluntaries (23 women), with a mean age was 33.6 (SD=8, range=20-50) and 16.5 (SD=3.5) years of formal schooling. The assessment of the 32 adults was performed by ALC and discussed with a clinical neuropsychologist (JJdP) and a psychiatrist (MAR-S).

Descriptive data were computed using the mean as a central tendency measure from the sample. Standard deviation, minimum and maximum values were also calculated. Normative values were defined using percentile scores and to assess the AQ reliability the Cronbach ‘s alpha was calculated.

## RESULTS

Based on AQ responses, we computed descriptive parameters using standard scores and percentiles following the original (Baron-Cohen et al., 2014) and Brazilian-adapted (Egito et al., 2018) scoring systems. AQ scores showed a normal distribution according to histogram analysis, and the mean score was 20.9 (SD=8.8).

We found good internal consistency in both genders (α=0,85 and 0.87), which means that the items reliably measured the same construct. Of the total of 91 men in our first sample, voluntaries with 32 points or more (the cut-off proposed by Baron-Cohen and colleagues) scored higher than approximately 93% of the control sample. Of 294 women, those who obtained 32 points or more, scored higher than approximately 97% of our sample (Table 1). In the original study, 32 points represented the 98th percentile (computed from the mean and standard deviations reported in the original paper). The study conducted by Osorio (Egito et al., 2018) examined the factor structure of the Brazilian version of the scale, and they proposed a three-factor model instead of five, a reduced version (25 items) and a different way to correct it. Besides that, they did not indicate a different cut-off for their sample. The distribution of AQ scores according to this method is shown in Table 1. In the second part of the study, based on the 32 score cutoff, all 32 adults got a clinical score on the AQ scale, but 24 also had a previous autism diagnosis. Two of them did not present enough characteristics to receive an autism diagnosis, but showed symptoms of other mental disorders (social phobia and generalized anxiety). Six volunteers of this group were classified as non-clinical although they showed borderline scores in AQ.

**Table 1.** Descriptive data of Autism Quotient scores stratified by scoring method and sex

|  | Cohen et al. (2001) |  | Egito et al. (2018) |  |
| --- | --- | --- | --- | --- |
|  | Male (N=91) | Female (N=294) | Male (N=91) | Female (N=294) |
| Mean | 25 | 20 | 62 | 54 |
| Std. Dev. | 8 | 9 | 12 | 13 |
| Min | 5 | 3 | 40 | 28 |
| Max | 43 | 45 | 86 | 94 |
| Pc.5 | 5 | 5 | 40 | 38 |
| Pc.10 | 10 | 7 | 44 | 41 |
| Pc.20 | 14 | 11 | 47 | 44 |
| Pc.30 | 16 | 13 | 50 | 46 |
| Pc.40 | 18 | 15 | 52 | 48 |
| Pc.50 | 20 | 17 | 54 | 50 |
| Pc.60 | 22 | 19 | 56 | 52 |
| Pc.70 | 24 | 21 | 58 | 54 |
| Pc.80 | 27 | 23 | 60 | 52 |
| Pc.90 | 30 | 27 | 64 | 59 |
| Pc.95 | 34 | 30 | 66 | 62 |
| Reliability | **0.87** | **0.85** | **0.76** | **0.82** |

Pc.: Percentile

## DISCUSSION

The Autism Spectrum Disorder was considered a diagnostic category, until the last edition of DSM have proposed a dimensional view for the clinical condition. ASD varies according to symptoms, severity, and need for support. It is expected that any population present from subclinical traits of ASD to very severe condition.

As seen in other populations AQ scores were normally distributed in our sample, and people with autism showed very high scores in the questionnaire (above the 90^th^ percentile). AQ also showed good reliability, both using the original (Baron-Cohen et al., 2001) and adapted (Egito et al., 2018) scoring systems. In our second sample, which showed scores above the cutoff score or reported a previous diagnosis of autism, we found 2 false-positive and 6 false-negative cases.

The availability of AQ as a screening instrument for autism traits in adults have huge importance, especially in the diagnostic process of adults with fewer impairments and preserved intelligence. These cases could be a challenge for health professionals. Besides that, knowing the level of autism symptoms enables the clinician to predict impairments, to offer adequate support, and to provide better guidance to the family. It is important to keep in mind that the scale alone is not enough for the diagnosis, but still a very useful tool in the investigation.

Finally, our results suggest that the studied population have a different profile compared to the original study, with clinical scores occuring at a lower percentile in our sample, which means that probably Brazilian subjects report more symptoms than other populations. Despite this, we observed that those traits were distributed in the typical population. Future studies are required to adequate the use of Autism Spectrum Quotient in the Brazilian population, such as to define a cut-off score that will better consider its culture and peculiarities.

## Data Availability

We have no ethical permission to make the data available. We might share data under request.

## ACKNOWLEDGMENTS

Alves ALC received an scholarship from Conselho Nacional de Desenvolvimento Científico e Tecnológico (CNPq). JJdP was the recipiente of a CAPES fellowship. DMM and MAR-S are CNPq research fellows. Finantial support from CNPq and FAPEMIG.

